# Acute Flaccid Paralysis Surveillance System Evaluation, Ho Municipality, Volta Region, Ghana, 2022

**DOI:** 10.1101/2024.12.07.24318650

**Authors:** Bakalilu Kijera, Abdul Nasir Alhasan, Sarja Jarjusey, Mary Bobb, Lamin F Manjang, Donne Ameme, Charles Lwanga Noora, Delia Bandoh, Ernest Kenu

**Affiliations:** Ghana Field Epidemiology and Laboratory Training Programme, School of Public Health, University of Ghana, Legon, Ghana; Ghana Health Services

**Keywords:** Acute Flaccid Paralysis, surveillance system, evaluation Ho Municipality, Ghana

## Abstract

**Background:** Ghana reported the last native Wild Poliovirus that causes poliomyelitis in 1999. However, Ghana experienced another outbreak when cases were imported into the country in 2003 and 2004. The decade-long polio-free status ended in 2019 following the detection of circulatory Vaccine-Derived Polio Virus type 2 (cVDPV2) through Environmental Health Surveillance (EHS). Ghana is currently free from cVDPV2 but is at risk of potential outbreaks. This evaluation describes the usefulness and attributes of an Acute Flaccid Paralysis (AFP) surveillance system and whether the system meets its objectives.

**Methods:** A descriptive evaluation of the AFP surveillance system from January 2017 to December 2021 was conducted in Ho Municipality, Volta Region. Interviews of health workers and key stakeholders, observations, records review, and administration of semi-structured questionnaires were made. The Centres for Disease Control and Prevention Updated guidelines for evaluating public health surveillance systems were used as a reference for developing a checklist. The quantitative data was analyzed using summary statistics and presented in percentages and frequencies. The qualitative interviews were audio recorded, transcribed, and narratively analyzed.

**Results:** Ten AFP cases were reported from January 2017 to December 2021. Six of these (28.57%) were detected based on actions taken on AFP surveillance data. The average NP-AFP was 2.14/100,000 with a minimum of 1.09/100,000 and a maximum of 4.12/100,000 under 15 population. One (10.0%) of the specimens reached the reference lab within 72 hours and an overall completeness of 68.04%. Three (30.0%) cases were follow-up after 60 days from the onset of the paralysis.

**Conclusion:** The AFP surveillance system in Ho municipality is partially meeting its objectives. It is useful, partially stable, and sensitive. The system is acceptable, representative, flexible, and accurate but poor in timeliness. The District Health Management Team should ensure specimens are sent to the reference laboratory within 72 hours after the case investigation.

## Introduction

Poliomyelitis is a contagious disease caused by the poliovirus which often affects children under 15 years of age (1). Polio is diagnosed with certainty when the poliovirus is isolated from a patient’s stools (2). Poliovirus can spread from person to person via the fecal-oral route and cause paralysis once it infects a person’s spinal code. (3).

There were 245 Wild Polio Virus Type1 (WPV1) isolated in 2020 globally and this has reduced significantly to 6 in 2021 (5). All the cases isolated in 2020 were reported in Eastern Mediterranean Region and the region reported 5 cases of the 6 cases reported in 2021. including 5 in 2021. Failure to eradicate polio in the remaining affected countries could result in a global resurgence of the disease (4). Circulatory Vaccine Derived Polio Virus Type 2 (cVDPV2) was also isolated (1,067 in 2020 and 659 in 2021) across the globe. In 2021, one (1) case of WPV1 was isolated in 2021 in the African Region (Malawi) and 530 and 521 of cVDPV2 were also isolated in 2020 and 2021 respectively (5).

In 1999, Ghana reported the last indigenous WPV case that causes poliomyelitis. However, 8 cases were imported into Ghana that caused an outbreak in 2003 and 2004 (6). Ghana’s decade-long polio-free status came to an end in 2019 following the detection of cVDPV2. The cVDPV2 outbreaks can occur in settings with low poliovirus population immunity and causes paralysis (7). There were 12 cases of cVDPV2 reported in 2020 and an overall total of 31 since the outbreak was declared in 2019 through Environmental Health Surveillance (EHS) (5).

Polio only affects humans, an effective vaccine is available and immunity is life-long (8). Maintaining very high immunization coverage and enhanced AFP surveillance has helped countries in the WHO African Region (9). Inactivated Poliovirus Vaccine (IPV) and three doses of Oral Poliovirus Vaccine (OPV) are part of the routine immunization schedule of Ghana (5). However, Acute Flaccid Paralysis (AFP) surveillance is the gold standard for detecting cases of poliomyelitis.

AFP surveillance involves the detection of enterovirus from the stool of children under the age of 15 years (10). Sensitivity of surveillance, completeness of reporting, case investigation and follow-up, and laboratory performance are the key indicators for AFP surveillance (11). The reasons for certification of polio eradication which is done on a regional basis are influenced by the quality of AFP surveillance across all countries in the region with at least no Wild Polio Virus (WPV) for 3 consecutive years (12).

In contrast, polio environmental surveillance is the routine collection and examination of sewage samples to detect the presence of the virus. Environmental surveillance can identify poliovirus circulation even in the absence of proven paralytic polio cases because paralysis only develops in 1% of poliovirus infections (13).

The AFP surveillance system in Ghana aims to detect cases of acute flaccid paralysis (AFP) and obtain laboratory confirmation of the aetiology of all suspected cases, report immediately all AFP cases using a case-based reporting form and collect at least 2 stool specimens (24 hours apart) from each case within 14 days. According to WHO, Ghana is currently free from cVDPV2 but is at risk of potential outbreaks. Thus, the AFP surveillance system is critical to maintaining standard. There may be underreporting of cases due to a lack of awareness or access to healthcare facilities in certain parts of Ho municipality. We evaluated the AFP surveillance system in Ho municipality to determine if the objectives are being met and assess the usefulness and attributes.

## Methods

### Evaluation Design

The evaluation of the surveillance system was done for the period January 2017 to December 2021. The CDC’s Updated Technical Guidelines for evaluating public health surveillance systems 2001, served as a guide in developing a semi-structured questionnaire/checklist for the evaluation. We interviewed stakeholders on the performance, usefulness, and attributes of the AFP surveillance system. We also reviewed AFP case-based forms for the period to determine the timeliness and completeness of reports submitted. Observations were made to verify key components of AFP surveillance in the municipality. The quantitative data were analyzed using percentages and frequencies while qualitative data were recorded and later transcribed to narrate the AFP surveillance system and its procedures.

### Evaluation Setting

The evaluation was conducted in Ho Municipality, one of the 18 districts of the Volta Region, Ghana. The municipality has five sub-districts (Hokpeta, Norvisi, Dutasor, Sokode/Akrofu, and Ho Central) and 48 health facilities and two hospitals. All the facilities in the district offer routine immunization services. Ho municipality is bordered northwest by the Ho West District, east and southeast by the Adaklu-Anyigba District, and southwest by Togo.

Ho Municipality has a total population of 184,209 and an under-fifteen (15) population of 96,266. The regional hospital is located in the district where patients are received from the boarder villages of Togo to seek medical care.

### Operation of the system

Once an AFP case is detected, two stool specimens are collected between 24 – 48 and within 14 days from the onset of paralysis. The specimens are packaged using the standard triple packaging system and sent to the WHO Polio reference lab within 72 hours after the investigation of the case. Feedback is expected within 21 days to the National AFP focal person and transcends the information to the region and then to the district for action if any. The stool adequacy, Sabin, and the Non-Polio Enterovirus are analyzed and a final classification is made. At 60 days a follow-up is made to understand whether it is residual paralysis or non-residual paralysis (fig 2).

**Fig 1:**
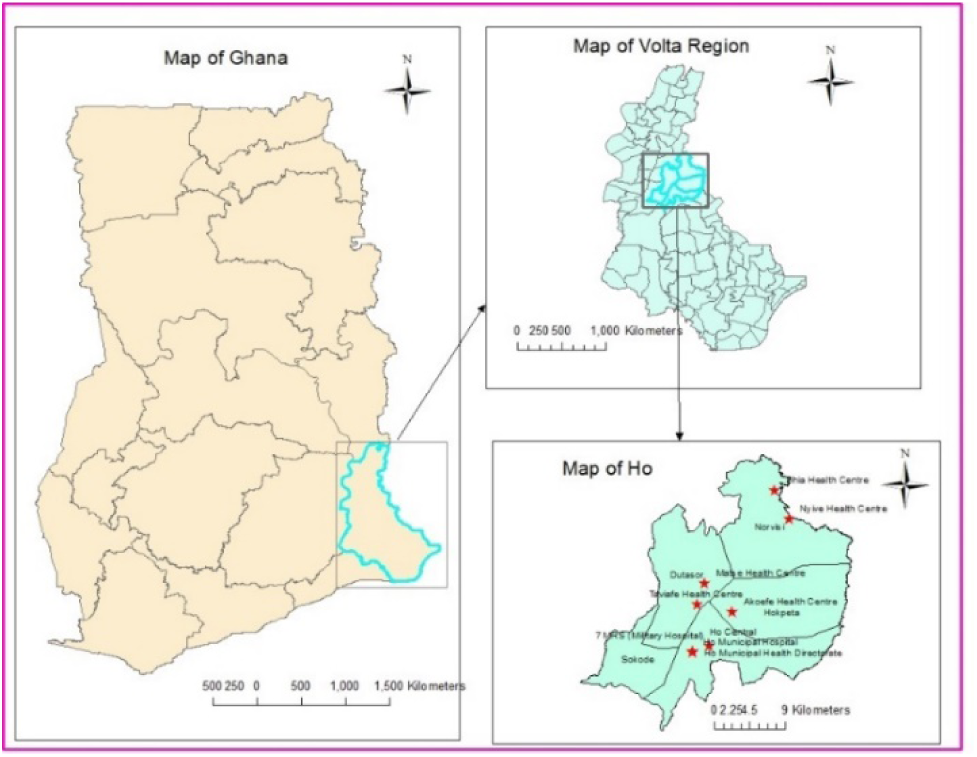
Evaluation Area Map, Ho Municipality.

**Fig 2:**
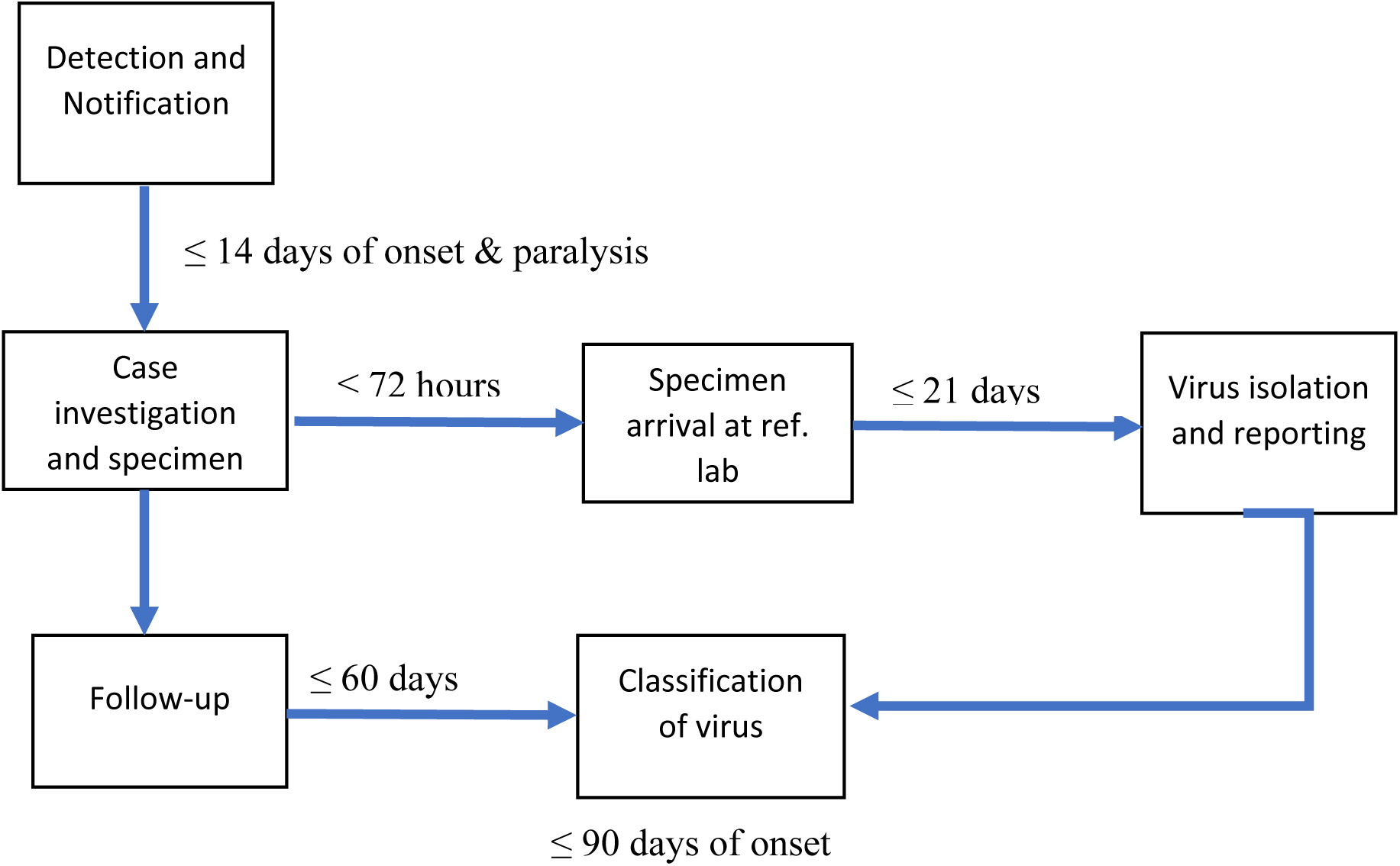
Operations of AFP surveillance system, Ghana.

Fig 3 depicts reporting and feedback on AFP surveillance in Ghana. All the levels from the community to the AFP focal person at the national disease surveillance are involved. The Disease Control Officer sends specimens to the reference laboratory and the results are shared with the AFP focal person at a national level.

**Fig 3:**
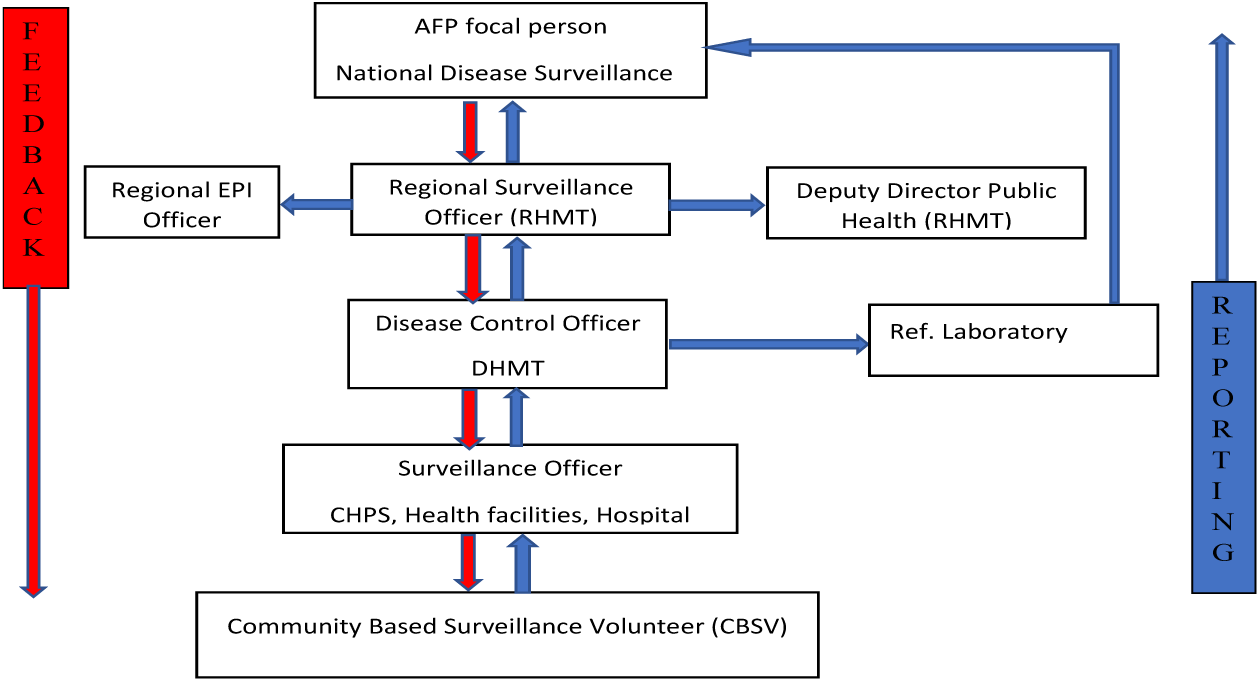
Algorithm of AFP surveillance reporting, Ho Municipality, Volta Region.

### Data Collection

We randomly selected six health facilities and one hospital in the AFP surveillance evaluation. We interviewed staff from the region, district, and health facilities from the 16^th^ to 30^th^ June 2022. We administered structured questionnaires and had an interview guide for the key informants.

We reviewed and observed surveillance tools and verified AFP surveillance indicators in all the facilities visited from the 6^th^ to 15^th^ June 2022. The collected data from key informants’ interviews were recorded using a mobile device and later transcribed.

### The system’s ability to meet its set objectives

The system’s ability to detect acute AFP cases and the availability of Case-Based Forms (CBF) of all the cases reported were assessed. The proportion of specimens of all suspected cases sent to the lab and the gap between the first specimen and the second specimen were also reviewed.

### The usefulness of the system (performance)

To assess the usefulness, we considered what data generated from the system is used for and how data generated from the system influenced AFP control measures in the municipality and country at large.

### Assessment of System Attributes

#### Simplicity

The easiness of the AFP case definition, the ease filling of out the Case-Based Form (CBF), and the operations of the AFP surveillance system were assessed to determine the simplicity.

#### Flexibility

We assessed what has changed in the case definition and case-based form, and whether these changes have affected or interrupted the smooth operations of the system.

#### Data quality

Data were reviewed to determine the completeness of AFP data from 2017 to 2021. The data generated in the health facility, district, region, and DHIMS2 were compared based on the evaluation period. At least ≥90% of data should be complete and reported on time.

#### Timeliness

The time duration between two specimens collected, investigated, and reported to the next level were reviewed from filled case-based forms for the period of evaluation.

The timeliness of AFP specimens to the WHO-accredited reference laboratory and the turn-around time were assessed.

#### Acceptability

Health facility staff were interviewed to assess their willingness to continue the AFP Surveillance System and their consistency in reporting weekly surveillance data. Community participation and the time surveillance officers are involved in AFP surveillance were also assessed. specimens arriving in good condition, and the proportion of AFP cases that were followed after 60 days of onset were also assessed.

#### Sensitivity

The Non-Polio Acute Flaccid Paralysis (NP-AFP) rate and stool adequacy were assessed to determine the system’s sensitivity.

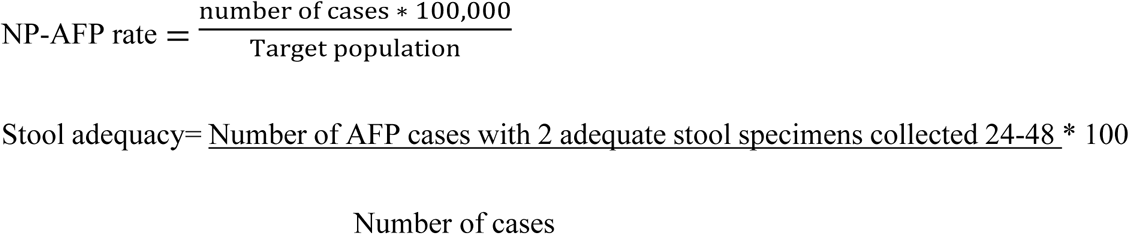

#### Representativeness

We evaluated the participation of health facilities in AFP surveillance and reviewed reported cases by sex, age, and place.

#### Stability

Assessment was done to understand whether there were interruptions of services due to power failure and unavailability of computers/tablets. Regional and district staff were interviewed to determine the human resources component as per the staffing norms.

### Stakeholders interviewed

We interviewed key informants/stakeholders of the system at the District Health Directorate, Regional Health Directorate (Regional Surveillance Officer), and staff from health facilities within the municipality. The Regional Surveillance Officer, District Surveillance Officer, and District Public Health Nurse were purposely selected as key informants.

### Acute Flaccid Paralysis Surveillance Case Definition

#### Suspected Case

A suspected case is defined as any child under 15 years of age with acute flaccid paralysis or any person with paralytic illness at any age in whom the clinician suspects poliomyelitis” (14).

#### Confirmed Case

A confirmed case is defined as a suspected case with a positive laboratory result (virus isolated in the stool) (14).

### Data Analysis

Quantitative data were analysed using frequencies and tables. Non-polio AFP rate and stool adequacy were analysed in rates and proportion respectively. Themes were developed to narrate the AFP surveillance system and its procedures. We showed the operations and reporting flow of AFP surveillance in Ghana.

### Ethical Statement

The Director of the Diseases Surveillance Department of the Ghana Health Service granted permission to conduct the evaluation. Permission was obtained from health facilities before the start of the assessment. All respondents were provided with the information sheet and written consent and were assured of confidentiality. Data was kept on password-protected devices and available only on a need-to-know basis. The authors had no access to information that could identify individual during or after the review.

## Results

A total of 5 health centers, 2 CHPS compounds, and 1 hospital were visited. The evaluation recruited 21 health workers amongst which, more than half of the respondents 52.38% (11/21) were nurses and 28.57% (6/21) were Disease Control Officers. The majority 61.9% (13/21) were females and most of the participants 57.14% (12/21) were between the ages of 20 – 30 years. The mean age of the participants was 31.9 ±6.5 years. (Table 1).

**Table 1:** Demographic characteristics of study participants.

| Characteristics | Frequency (%) |
| --- | --- |
| **Sex** |  |
| Male | 8 (38.1) |
| Female | 13 (61.9) |
| **Age** |  |
| 20-30 | 12 (57.14) |
| 31-40 | 8 (38.10) |
| Above 40 | 1 (4.76) |
| **Designation** |  |
| Disease Control Officer | 6 (28.57) |
| Nurse | 11 (52.38) |
| Physician Assistant | 1 (4.76) |
| Filed assistant | 2 (9.52) |
| Public Health Nurse | 1 (4.76) |

### The system’s ability to meet its set objectives

During the five years, the district reported at least 1 case each year – albeit they have not met the NP-AFP rate of 2/100,000 in 2018 and 2019. The district recorded 1.11/100,000 and 1.09/100,000 populations in 2018 and 2019 respectively. However, the laboratory results were not available in the district and feedback comes in 2 to 6 months after the sample is taken to the laboratory. The case-based forms of all the cases reported were available at the district.

All the cases were investigated and notified immediately. Two stool specimens were collected within 24 - 48 hours apart from all the AFP cases reported during the evaluation period.

### The usefulness of the system (performance)

The region and the district monitor the trend graph of AFP cases and use the data to make informed decisions. However, they had a meeting but no minutes were available. It was highlighted that in the last quarter of 2021, the municipality embarked on an active case search funded by WHO to meet the NP-AFP rate district target.

### Attributes of the AFP surveillance system

#### Simplicity

Most of the participants 13 (61.9%) knew the case definition and 4 (19.50%) had one time identified a case and completed the case-based form. It takes 21 days before the polio virus is isolated and reported.

#### Flexibility

The AFP reporting form was modified from 44 variables to 75 variables. Among the respondents, 80.95% (17/21) said, the changes have not disrupted the smooth operations of the surveillance system. More than three-quarters 76.10% (19/21) of the respondents had used the electronic database to report.

#### Data Quality

The number of AFP cases reported by the district and the data shown in the region and the DHIMS2 were 100% accurate. However, the overall percentage completeness of the variables filled in the CBF reported from 2017 to 2021 was 68.04%.

#### Timeliness

All the reported cases were investigated on time and stool specimens from the cases were collected 24-48 hours apart and within 14 days. There were 90% (9/10) of the specimens sent to the reference lab after 72 hours of collection.

#### Sensitivity

The non-Polio AFP rate per 100,000 was less than the targets in 2018 (1.11/100,000), 2019 (1.09/100,000), and 2020 (2.12/100,000).

The average NP-AFP was 2.14/100,000 with a minimum of 1.09/100,000 and a maximum of 4.12/100,000 under 15 population. The Non-polio enterovirus (NPENT) was 0% from 2018 20 2022. Overall, only 20% (2/10) had Non-polio enterovirus in the stool. (Table 2).

**Table 2:** Acute Flaccid Paralysis Surveillance Performance Indicator, Ho Municipality, 2017-2021.

| Indicators of surveillance performance | Target | District performance |  |  |  |  |
| --- | --- | --- | --- | --- | --- | --- |
|  |  | 2017 | 2018 | 2019 | 2020 | 2021 |
| Number of AFP cases investigated | 2 | 2 | 1 | 1 | 2 | 4 |
| Annualized Non-polio AFP rate per 100,000 children $\leq 15$ years | $\geq 2$ | 2.28 | 1.11 | 1.09 | 2.12 | 4.12 |
| NPENT | 10% | 50% | 0% | 0% | 0% | 25% |
| Proportion of AFP cases investigated within 48 hours | $\geq 80\%$ | 100 | 100 | 100 | 100 | 100 |
| Proportion of AFP cases with 2 adequate stool specimens collected 24-48 hours apart and $\leq 14$ days after onset | $\geq 80\%$ | 100 | 100 | 100 | 100 | 100 |
| Proportion of specimens arriving at the laboratory in good conditions | $\geq 80\%$ | 100 | 100 | 100 | 100 | 75 |
| Proportion of specimens arriving at WHO accredited lab within 72 hours | $\geq 80\%$ | 0 | 0 | 0 | 0 | 25 |
| Proportion of specimens for which laboratory results were sent within 21 days | $\geq 80\%$ | 0 | 0 | 0 | 0 | 0 |
| Proportion of AFP cases that were followed 60 days after the onset of paralysis | $\geq 80\%$ | 100 | 0 | 0 | 50 | 0 |

#### Acceptability

All the facilities visited are willing to continue AFP surveillance – albeit more than half of the respondents 12 (57.14%) lack support in their day-to-day activities. All 8 facilities report weekly and recorded 100% timeliness were 100%. The proportion of specimens arriving at the laboratory in good condition was 100% across the five years except in 2021 which recorded 75%.

All the AFP cases reported were investigated within 48 hours but only one specimen arrived at the reference laboratory within 72 hours. All the cases were discarded and there were only three cases that were followed (2 cases in 2017 and 1 case in 2020) 60 days after the onset of the paralysis.

One of the stakeholders highlighted that “*staff lack motivation to partake in surveillance and most of them do not read books to broaden their scope”.* Community participation in the urban areas were poor as opposed to the rural communities. Almost all participants 20 (95.24%) believe that community participation is moderate.

#### Representativeness

All the health facilities in the district participate in AFP surveillance. Cases were reported from both males (4) and females (6), and 70% were under five years. However, the 10 reported cases for the five years came from only two sub-districts i.e., Norvisi and Ho Central.

#### Stability

“*High staff attrition and inadequate specimen collection tools is a challenge in Ho Municipality”* (Staff RHMT). All the facilities visited have either functional computers or tablets to enter routine surveillance data in the DHIMS2. All the facilities visited displayed a copy of the case definition at the outpatient department.

## Discussion

This study evaluated the AFP surveillance system in Ho municipality to determine if the objectives are being met and assess the attributes and usefulness. We found that two stool specimens were collected within 24 - 48 hours apart and within 14 days from all the AFP cases reported during the evaluation period. Our study also provides evidence that feedback on the laboratory results of the cases was never available within the expected 21-day period. In addition, three cases were followed 60 days after the onset of the paralysis, and one AFP case was sent on time (within 72 hours) to the reference laboratory.

The Volta Region met the NP-AFP rates per 100,000 targets in 2018 and 2019 (15) but the Ho Municipality did not meet the target. A similar finding was observed in Kenya where progress has been made at the national level but a declining trend in some other regions (16). The average NP-AFP rate was lower than the study in Northern Sudan which reported 4/100,000 under-15 population (17). We also found that the NP-AFP rate was 1.09/100,000 to 4.12/100,000. The findings are comparable to the study done in South Sudan with a minimum and maximum rate of 1.2 to 4.4 per 100,000 population respectively. High staff attrition and inadequate specimen collection tools have compromised the stability of the system. Different outbreak settings (regional vs districts) could be the reasons for the disparity. Staff that have good knowledge of AFP are in the district and rarely do they have direct contact with patients.

We found that the surveillance system was useful and the district staff were entirely responsible for monitoring trends. However, meeting minutes were not available in the district health office as part of the discussion to make decisions and take some actions on AFP surveillance. Similar study findings have been observed in Mashonaland West Province (1) where the surveillance system was perceived useful and health workers are more likely to participate diligently. Failure to produce meeting minutes purportedly is an indication that the meeting was probably not held.

In addition, the findings showed that the AFP surveillance system was complex and all the facilities visited had the case definition displayed on the wall in the OPD department. These findings were not in conformity with a study in Nigeria which reported that the AFP surveillance system is simple due to the ease of filling out forms and the case definition was simple too (18). This could be explained as all the health facilities visited did not have a single empty form of AFP CBF and had no idea how to fill out the form.

All specimens were collected within 24-48 hours but specimens were not transported timely to the reference laboratory. A similar finding was reported in Mwenezi district, Masvingo (19). This could be because of the distance from the laboratory and the logistics involved in sending the specimens. A delay in sending specimens can reduce the chances of isolating the virus.

The data found in the district were consistent with what was shown at the regional level. The data completeness was lower compared to 97% completeness of a study conducted in Kebbe State, Nigeria. Lack of feedback on laboratory results of cases to complete the outcome of the AFP case on the CBF could be the reason for the lower completeness of the data. The date specimen sent to the lab is often missing in some of the cases reported. This could also mean that the specimen was not sent to a laboratory for processing.

We found that the sex distribution of AFP cases was higher among females. The finding is not in conformity with a study in Jordan and Sudan that found that 55.8% and 51% of cases were males respectively (17, 18). However, the differences were minute and all sexes have equal exposures to AFP. Most of the cases were under five years and do not conform to the study in Sotoko State, Nigeria (20) where 63% of cases were between 10-15 years old. Predictive value positive (PVP) is defined as “the proportion of reported cases that have the health-related event under surveillance” (23). It is difficult to determine PVP for AFP surveillance in Ghana as the diagnosis is strictly clinical and only the true AFP cases are reported.

## Conclusion

The AFP surveillance system in Ho municipality is partially meeting its objectives. It is useful, partially stable, and partially sensitive. The system is acceptable, representative, and flexible but poor in data quality and timeliness. The evaluators explained the AFP case definition to staff at the outpatient department using what was displayed on the wall. We also demonstrated the monitoring trend of weekly surveillance data to the health facility surveillance officers. We also conducted a debriefing at the District Health Management Team (DHMT) on the findings and the way forward. We recommend that the district should follow up on laboratory results of cases, and ensure specimens are sent to the reference laboratory within 72 hours after the case investigation and strengthen the 60-day flow-up of all AFP cases.

## Data Availability

All relevant data are within the manuscript and its Supporting Information files.

file:///C:/Users/bkkij/Downloads/S3%20File.%20Data%20set.htm

## Acknowledgment

I acknowledge Ghana Field Epidemiology and Laboratory Training Program and Ghana Health Services for their immense support during the one-day field one. Special Thanks go to the Regional Health Directorate Volta Region and District Health Management of Ho Municipality, Ghana, for given us permission to conduct the evaluation in Ho District.

## Supporting document

S1_File. AFP cases S2_File. Questionnaires S3+File. Dataset

